# Preferences of people living with HIV for features of tuberculosis preventive treatment regimens – a discrete choice experiment

**DOI:** 10.1101/2023.09.13.23295043

**Authors:** Hélène E. Aschmann, Allan Musinguzi, Jillian L. Kadota, Catherine Namale, Juliet Kakeeto, Jane Nakimuli, Lydia Akello, Fred Welishe, Anne Nakitende, Christopher Berger, David W. Dowdy, Adithya Cattamanchi, Fred C. Semitala, Andrew D. Kerkhoff

## Abstract

**Background:** Tuberculosis (TB) preventive treatment (TPT) is recommended for people living with HIV (PLHIV) in high TB burden settings. While 6 months of daily isoniazid remains widely used, shorter regimens are now available. However, little is known about preferences of PLHIV for key features of TPT regimens.

**Methods:** We conducted a discrete choice experiment among adult PLHIV engaged in care at an urban HIV clinic in Kampala, Uganda. In nine random choice tasks, participants chose between two hypothetical TPT regimens with different features (pills per dose, frequency, duration, need for adjusted antiretroviral therapy [ART] dosage and side effects). We analyzed preferences using hierarchical Bayesian estimation, latent class analysis, and willingness-to-trade simulations.

**Results:** Of 400 PLHIV, 392 (median age 44, 72% female, 91% TPT-experienced) had high quality choice task responses. Pills per dose was the most important attribute (relative importance 32.4%, 95% confidence interval [CI] 31.6 – 33.2), followed by frequency (20.5% [95% CI 19.7 – 21.3]), duration (19.5% [95% CI 18.6 – 20.5]), and need for ART dosage adjustment (18.2% [95% CI 17.2 – 19.2]). Latent class analysis identified three preference groups: one prioritized less frequent, weekly dosing (N=222; 57%); another was averse to ART dosage adjustment (N=107; 27%); and the last prioritized short and tolerable regimens (N=63; 16%). All groups highly valued fewer pills per dose. Participants were willing to accept a regimen of 2.8 months’ additional duration [95% CI: 2.4 – 3.2] to reduce pills per dose from five to one, 3.6 [95% CI 2.4 – 4.8] months for weekly rather than daily dosing, and 2.2 [95% CI 1.3 – 3.0] months to avoid ART dosage adjustment.

**Conclusions:** To align with preferences of PLHIV, decision-makers should prioritize the development and implementation of TPT regimens with fewer pills, less frequent dosing, and no need for ART dosage adjustment, rather than focus primarily on duration of treatment.

## Introduction

Tuberculosis (TB) preventive treatment (TPT) is strongly recommended to address the high disease burden among people living with HIV (PLHIV) in TB endemic settings [1]. Short-course TPT regimens have been shown to be similarly effective and better tolerated than the conventional 6 or 9 months of daily isoniazid (6H or 9H) and are now recommended as options for TPT in updated World Health Organization (WHO) guidelines [1]. These regimens include 3HP (three months of weekly isoniazid and rifapentine), 1HP (one month of daily isoniazid and rifapentine), 3HR (three months of daily isoniazid and rifampin), and 4R (four months of daily rifampin). However, data on the preferences of PLHIV for key features that comprise and differentiate each of these regimens – such as treatment duration, frequency of dosing, number of pills per dose – are lacking.

Current WHO guidelines on TB preventive treatment were informed by a single preference study including only 10 participants living with HIV [2]. Preferences were assessed using Likert-scale questions and participants rated all features evaluated as important (short duration, less frequent intake, fewer side-effects, fewer clinic visits, fewer pills, no need for directly observed therapy (DOT), and no need to change dosage of antiretroviral therapy [ART]). However, it remains unknown how PLHIV would value individual features, make trade-offs between features, and ultimately choose between TPT regimens with different features. Such data are critical to inform decisions on scaling up TPT regimens and to guide future TPT regimen development.

Choice-based preference elicitation methods, including discrete choice experiments (DCEs), are increasingly being utilized to more systematically characterize patients’ healthcare preferences and inform policy- and implementation-related decisions [3–6]. Compared to simple rating exercises such as Likert scale questions, DCEs measure trade-offs through a series of repeated questions where participants must choose between two or more hypothetical alternatives (e.g., “would you prefer option A or option B?”). Notably, DCEs have been shown to have good predictive value for health-related choices [7], including for TPT regimens in a low TB burden setting [8].

We therefore conducted a DCE among adult PLHIV accessing routine HIV care in Kampala, Uganda. Our objectives were to 1) determine the relative importance of TPT regimen features; 2) simulate how willing participants were to trade one TPT feature for another; and 3) assess the heterogeneity in preferences and identify distinct subgroups of PLHIV with similar preferences.

## Methods

### Setting and participants

We conducted a cross-sectional survey that included a DCE from July to November 2022. The study took place at the Mulago Immune Suppression Syndrome (i.e., HIV/AIDS) clinic, in Kampala, Uganda. The clinic provides comprehensive HIV care to over 16,000 PLHIV and is the largest outpatient HIV clinic in the country.

Individuals were eligible for study participation if they were receiving HIV/AIDS care at the clinic, were 18 years or older, had not initiated a TPT regimen in the past year, and were not currently receiving TB treatment. People who were unable or unwilling to provide informed consent or were currently incarcerated were excluded. We defined the inclusion criteria to include individuals eligible for initial TPT or likely to qualify for repeated TPT in the future.

### Ethics, consents, and procedures

The Makerere University School of Public Health Research and Ethics Committee, the University of San Francisco Institutional Review Board and the Uganda National Council for Science and Technology approved the study. All participants provided written informed consent.

### DCE design

The DCE was designed using the “balanced overlap” method in Sawtooth Lighthouse Studio version 9.13.2 [9] to allow for analysis of interaction terms [10]. An initial list of DCE attributes and levels reflecting key features of TPT regimens was generated based on a review of the literature, refined by an interdisciplinary team, and further refined after pilot testing among 29 PLHIV (Appendix A). The final design included six attributes with 3 levels each, except for “need to adjust dose of ART” which included 2 levels (Figure 1). Each participant was randomly allocated to one of 500 randomly generated sets of nine random choice tasks. In addition, we included a dominant choice task to assess participant comprehension of the DCE (Appendix Table 1). All choice tasks were unlabeled (named “Treatment A” and “Treatment B”). Participants were first required to choose their preferred treatment (A or B), and then between their preferred treatment and no treatment (Appendix Figure 1).

**Figure 1:**
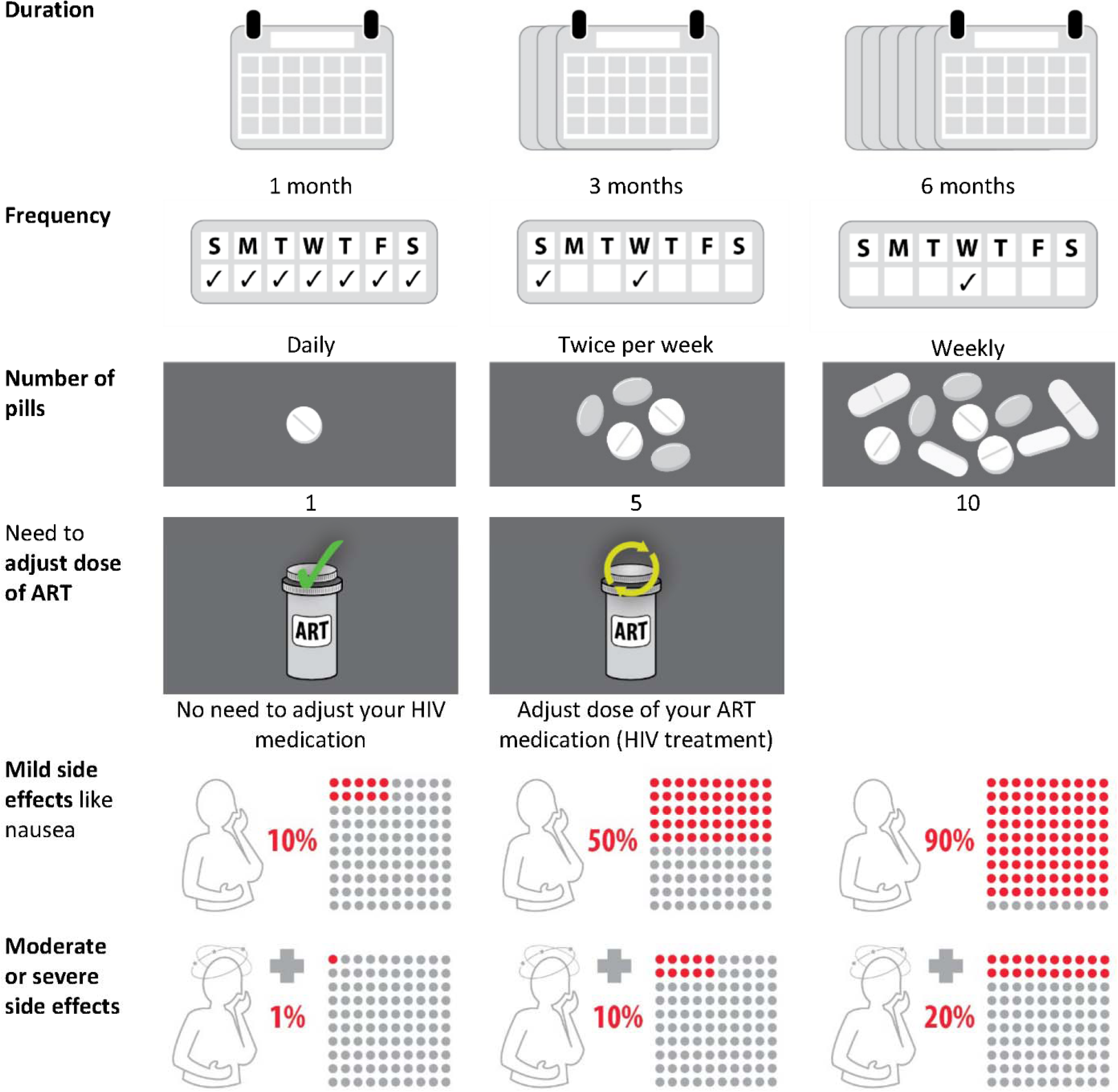
Attributes and levels in the discrete choice experiment describing different tuberculosis preventive treatment regimens. This figure shows the final selection of attributes (column 1) and how levels were depicted to participants (columns 2-4). ART: antiretroviral therapy, HIV: human immunodeficiency virus

### Procedures

The survey was administered one-on-one by a trained interviewer using an electronic tablet. Interviewers first presented general information on TB prevention using a flipbook. Interviewers then explained the DCE attributes and levels using a flipbook with icons from the DCE (Appendix B). In addition to the DCE component, the survey collected information on demographics, multidimensional poverty index [11], and TB/HIV history.

### Sample size

We considered the minimum sample size for the DCE to be 250 PLHIV based on the formula 500c/ta, where ‘c’ is the product of the greatest number of levels for any two attributes, ‘t’ is the number of choice tasks, and ‘a’ is the number of options per choice task [12]. To enable a pre-specified subgroup analysis by sex, we targeted a sample size of 400 participants since a minimum of 200 participants per subgroup is recommended [12].

### Statistical analysis

All analyses were performed in Lighthouse Studio version 9.13.2 (Sawtooth Software) and R version 4.1.2. We excluded participants from analyses if: (1) the dominant fixed choice task was answered incorrectly, (2) the no treatment option was always selected (indicating the participant was not interested in TPT), or (3) participants showed two or more signs of inattention or lack of understanding including ‘straight-lining’ (e.g., always choosing option A or option B), self-reported difficulty understanding the tasks (“difficult” or “very difficult”) in a question at the end of the survey, and inconsistent choices indicated by a root-likelihood (RLH) fit statistic below 0.651. The RLH threshold was selected based on simulations of random answers as described previously [13].

We used a hierarchical Bayesian model to calculate mean preference weights (also known as part-worth utilities) for each attribute level (along with 95% CIs) and the relative importance of attributes (which add up to 100% across attributes). We used latent class multinomial logit to identify groups of participants with distinct preferences and examined an elbow plot of the model fit criteria as well as average group membership probability to determine the number of latent class groups [14–16]. We implemented a Shares of Preference Model using the Sawtooth Choice Simulator tool to estimate participants’ willingness-to trade treatment duration (in months) and number of pills per dose for other regimen features using 3HP (4 fixed-dose combination pills weekly for 3 months with 11.5% mild and 6.0% moderate or severe side effects) and 6H (2 pills daily for 6 months with 36.1% mild and 8.2% moderate or severe side effects) as competitors given their current availability as TPT options in Uganda. We defined moderate or severe side effects as requiring a clinic visit, and used reported adverse event rates for the simulations [17]. We calculated 95% CIs using 300 bootstrap samples and 30 competitive sets per sample. We performed subgroup analyses according to preference groups identified from the latent class analysis.

## Results

### Participant characteristics

Of 456 persons screened, 414 were invited and 401 consented (response rate of 97% [401/414]) (Figure 2). We excluded one person from the study who was erroneously enrolled three times, as well as eight additional participants based on quality checks: three failed the dominant task, three had two signs of inattention or difficulty understanding choice tasks, and two indicated no interest in taking TPT (always selecting the no treatment option).

**Figure 2:**
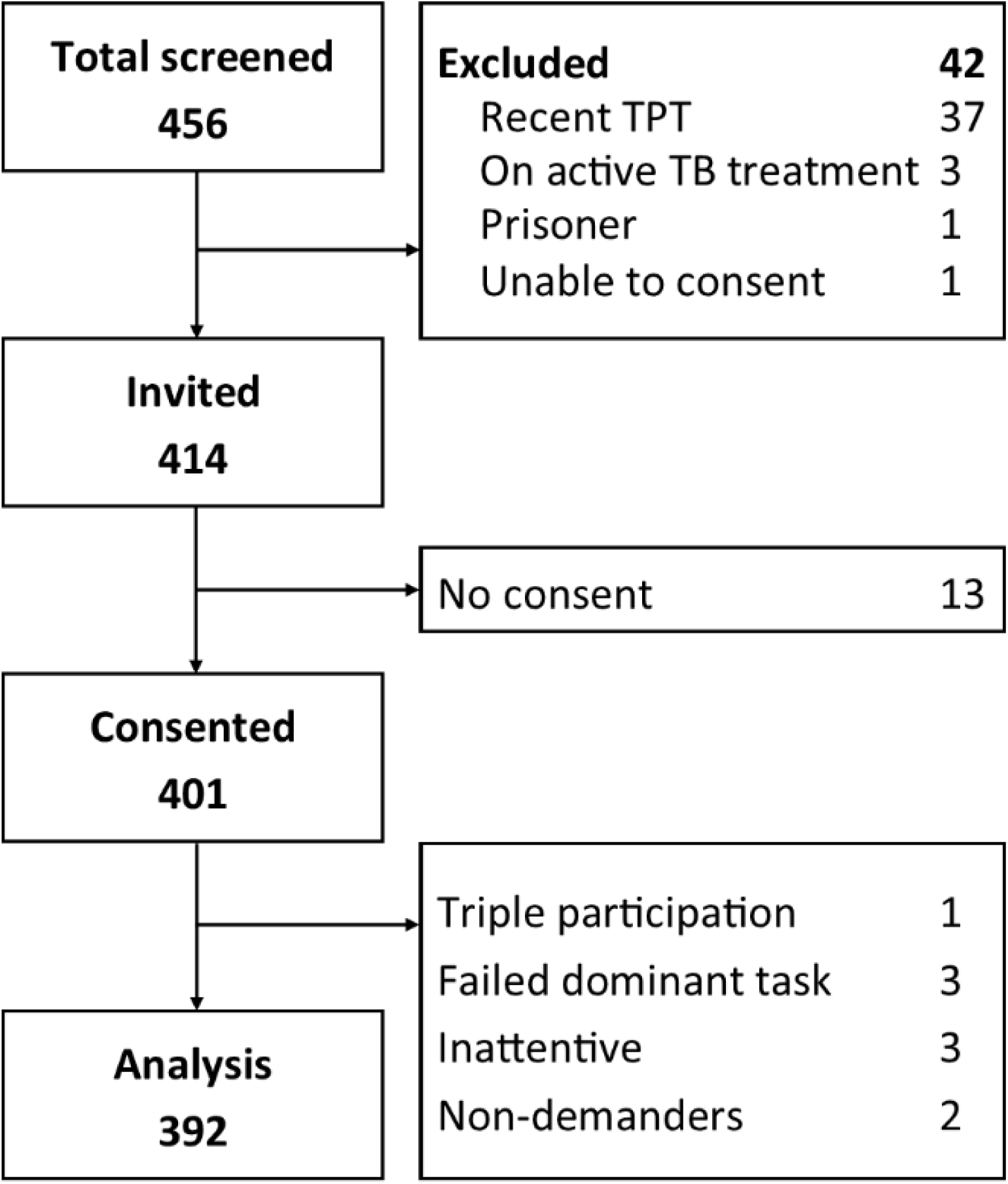
Participants’ study flow with 392 participants included in the final analysis.

The majority of participants were female (72%), employed (80%), and ART-experienced, with a median of 10.3 years on ART (Table 1). Most participants had previously taken TPT (91%), with either 6H (68%) and/or 3HP (33%). Most participants found it easy or very easy to understand the DCE (88%) and to choose between TPT options in each DCE choice task (77%). Non-participants (13 persons eligible and invited but who did not consent) were similar to participants with respect to age (mean 46.9, interquartile range (IQR): 41 to 54) and sex (77% female).

**Table 1:** Participant characteristics at baseline, based on medical records and self-report.

| Participants (N = 400) | N | (%) |
| --- | --- | --- |
|  | mean | (IQR) |
| **Female sex** | 288 | (72%) |
| **Age** | 44.3 | (IQR: 38, 51) |
| **BMI** | 27.0 | (IQR: 22.2, 30.1) |
| **Education** |  |  |
| None | 91 | (23%) |
| Primary | 150 | (38%) |
| Secondary | 116 | (29%) |
| Tertiary or higher | 43 | (11%) |
| **Work status** |  |  |
| Hired | 76 | (19%) |
| Self-employed | 245 | (61%) |
| Unemployed | 45 | (11%) |
| Not working | 22 | (6%) |
| Other | 12 | (3%) |
| **Multidimensional poverty index<sup>1</sup>** |  |  |
| Severely poor | 11 | (3%) |
| Poor | 63 | (16%) |
| Vulnerable | 111 | (28%) |
| **Prior TPT<sup>2</sup>** |  |  |
| Prior 6H | 247 | (68%) |
| Prior 3HP | 121 | (33%) |
| **TPT completion (N = 365)** |  |  |
| TPT completed | 353 | (97%) |
| Do not know / do not want to answer | 1 | (0%) |
| **Did you experience side effects from TPT? (N=365)** |  |  |
| Yes, and I had to see my doctor about it. | 28 | (8%) |
| Yes, but only mild ones and I did not see my doctor about it. | 58 | (16%) |
| **History of active tuberculosis** | 72 | (18%) |
| **Current antiretroviral therapy** |  |  |
| Dolutegravir-based | 378 | (95%) |
| Efavirenz-based | 14 | (4%) |
| Other | 8 | (2%) |
| **Time on antiretroviral therapy (years)** | 10.4 | (IQR: 7.2,14.1) |
| **Viral load** |  |  |
| Suppressed | 394 | (99%) |
| Unsuppressed ($\geq 1000$ copies) | 3 | (1%) |
| Not yet done, recent HIV diagnosis | 1 | (0%) |
| Missing | 2 | (1%) |
| **Taking other medications<sup>3</sup>** | 147 | (37%) |
| **Herbal medicine use** |  |  |
| Within last month | 90 | (23%) |
| Within last year | 98 | (24%) |
| Longer than a year ago | 128 | (32%) |
| **Hormonal contraceptives among women (N=288)** | 52 | (18%) |

1. The multidimensional poverty index captures deprivations in health, education, and living standards. 2. 3 persons answered that they had both previously taken 6H and 3HP. 3. Currently taking other medications not including HIV medication or contraceptives. 3HP: 3 months of isoniazid and rifapentine, 6H: 6 months of isoniazid, BMI: body mass index, HIV: human immunodeficiency virus, IQR: interquartile range, TPT: tuberculosis preventive therapy.

### Preferences for TPT regimen features

Overall, participants assigned the highest relative importance to the number of pills per dose (32.4% [95% CI 31.6 – 33.2]), with one pill per dose being strongly preferred compared to 10 pills per dose (Figure 3, Appendix Table 2). Frequency of TPT dosing (relative importance 20.5% [95% CI 19.7 – 21.3]), duration of TPT (relative importance 19.5% [95% CI 18.6 – 20.5]), and need for ART dosage adjustment (relative importance 18.2% [95% CI 17.2 – 19.2]) were all similarly important. Weekly frequency was preferred over twice per week and daily dosing, 1-month duration was preferred compared to 3 or 6 months, and regimens not requiring ART dose adjustment were strongly preferred compared to those requiring ART dose adjustment (Figure 3). Side effects were considered much less important than other attributes (relative importance 5.0% [95% CI 4.6 – 5.4] for mild side effects and 4.4% [95% CI 4.1 – 4.7] for moderate or severe side effects).

**Figure 3:**
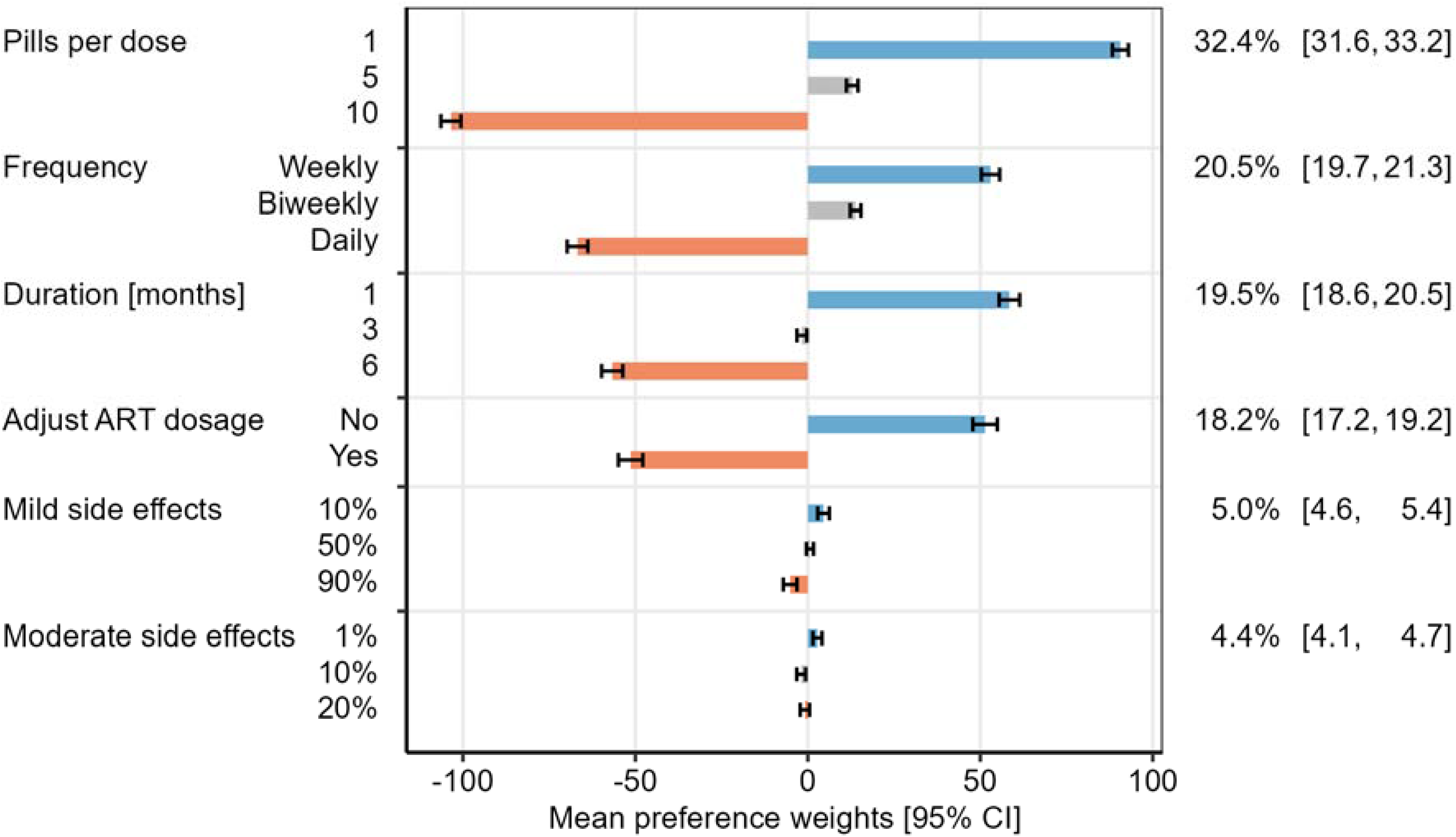
Mean preference weights of attribute levels and relative importance of attributes among all participants. Bars indicate the mean preference weights for each level among 392 participants using hierarchical Bayesian estimation. Blue bars indicate levels with the strongest positive preference (most preferred) per attribute, orange bars indicate levels with negative preference (least preferred). The percentage on the right side indicates the mean relative importance for each attribute.

### Heterogeneity of preferences for TPT features

Using latent class analysis, we identified three preference groups (Figure 4, Appendix Figure 2), all of whom preferred fewer pills per dose. The largest group (N=222, 57%) also prioritized less frequent dosing (“non-daily doses”). Another group (N=107, 27%) strongly preferred TPT regimens that required no ART dosage adjustment (“keep ART as is”). Finally, the last group (N=63, 16%) preferred shorter regimens with a lower risk of mild side effects (“short and tolerable”).

**Figure 4:**
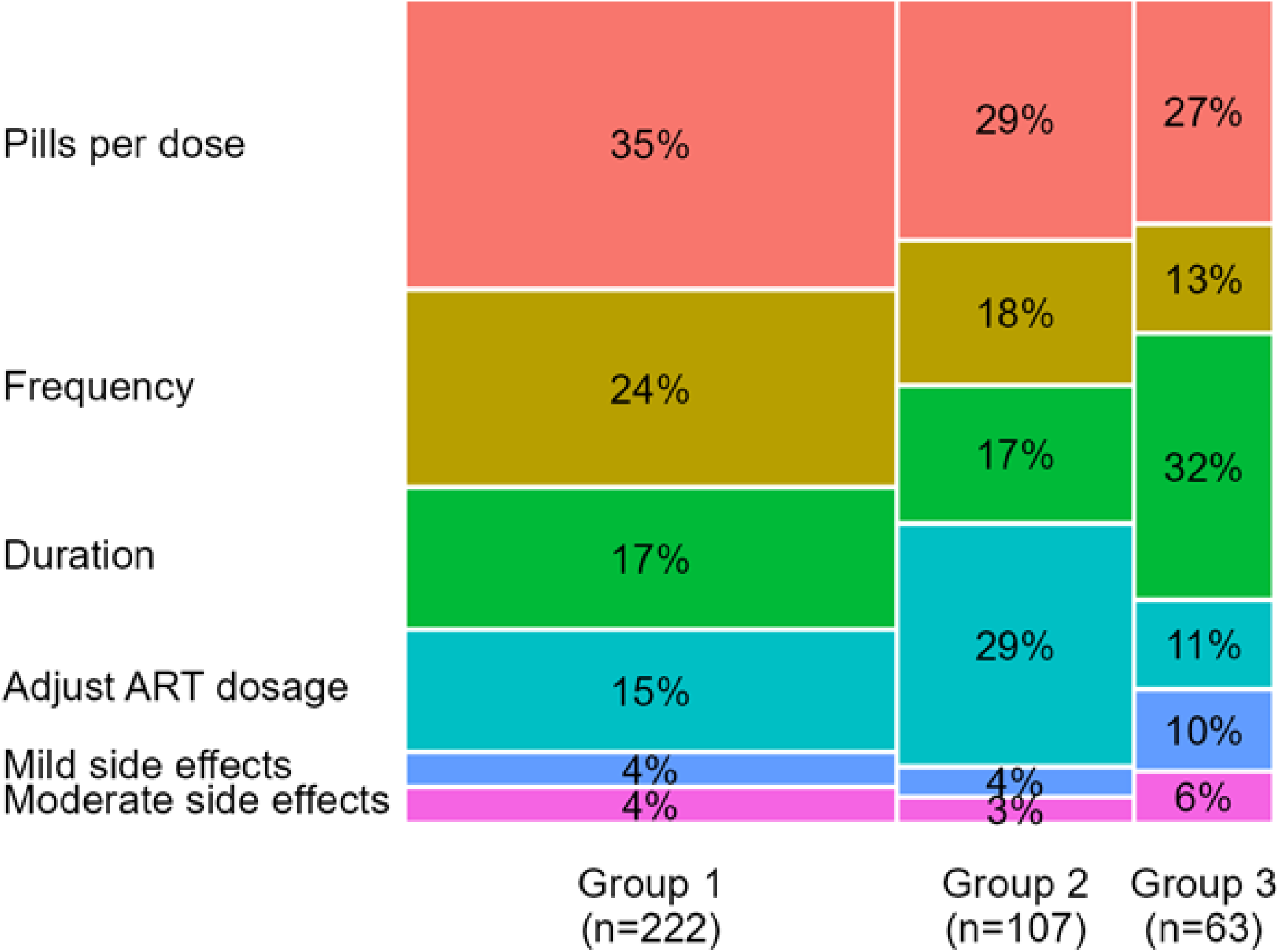
Mosaic plot showing the mean relative importance modeled using hierarchical Bayesian analysis among three groups identified by latent class analysis. The width of each column corresponds the proportion each group comprises of the overall population.

### Associations of preferences with baseline characteristics

We explored the association of baseline characteristics with latent class membership and individual preference weights. We found no association between sex, age, poverty, working status, prior history of TB, and years on ART with individual preference weights for duration, number of pills per dose, frequency of dosing and side effects (Appendix Table 3). However, participants with less ART experience were more averse (i.e., had stronger negative preferences) to TPT regimens requiring ART dosage adjustments (Appendix Table 3) and were more likely to be in the “keep ART as is” group (Appendix Table 4). In addition, participants taking other medications were more averse to ART dosage adjustments, and participants with any education were less averse to a high risk (90%) of mild side effects than participants with no education (Appendix Table 3).

### Willingness-to-trade for more preferred TPT regimen features

We simulated the tradeoff between treatment duration (in months) and other regimen features (Figure 5A). Participants were willing to take TPT for 2.7 [95% CI: 1.8 – 3.5] additional months in exchange for reducing the number of pills per dose from 10 to 5. If the number of pills per dose could be further reduced from 5 to 1, participants were willing to take TPT for another additional 2.8 [95% CI: 2.4 – 3.2] months. Participants were willing to take TPT for 3.6 [95% CI 2.4 – 4.8] additional months in exchange for weekly rather than daily dosing, and for 2.2 [95% CI 1.3 – 3.0] additional months in exchange for not needing ART dosage adjustment. Participants were willing to take TPT for only 0.6 [95% CI 0.3 – 0.9] additional months to reduce the risk of mild side effects from 90% to 10%, and were not willing to trade a longer duration of treatment for a lower risk of moderate or severe side effects. For all regimen features assessed except for moderate or severe side effects, willingness-to-trade varied between preference groups identified by latent class analysis (Appendix Figures 3 and 4). Corresponding trade-offs for the number of pills per dose are presented in Figure 5B.

**Figure 5:**
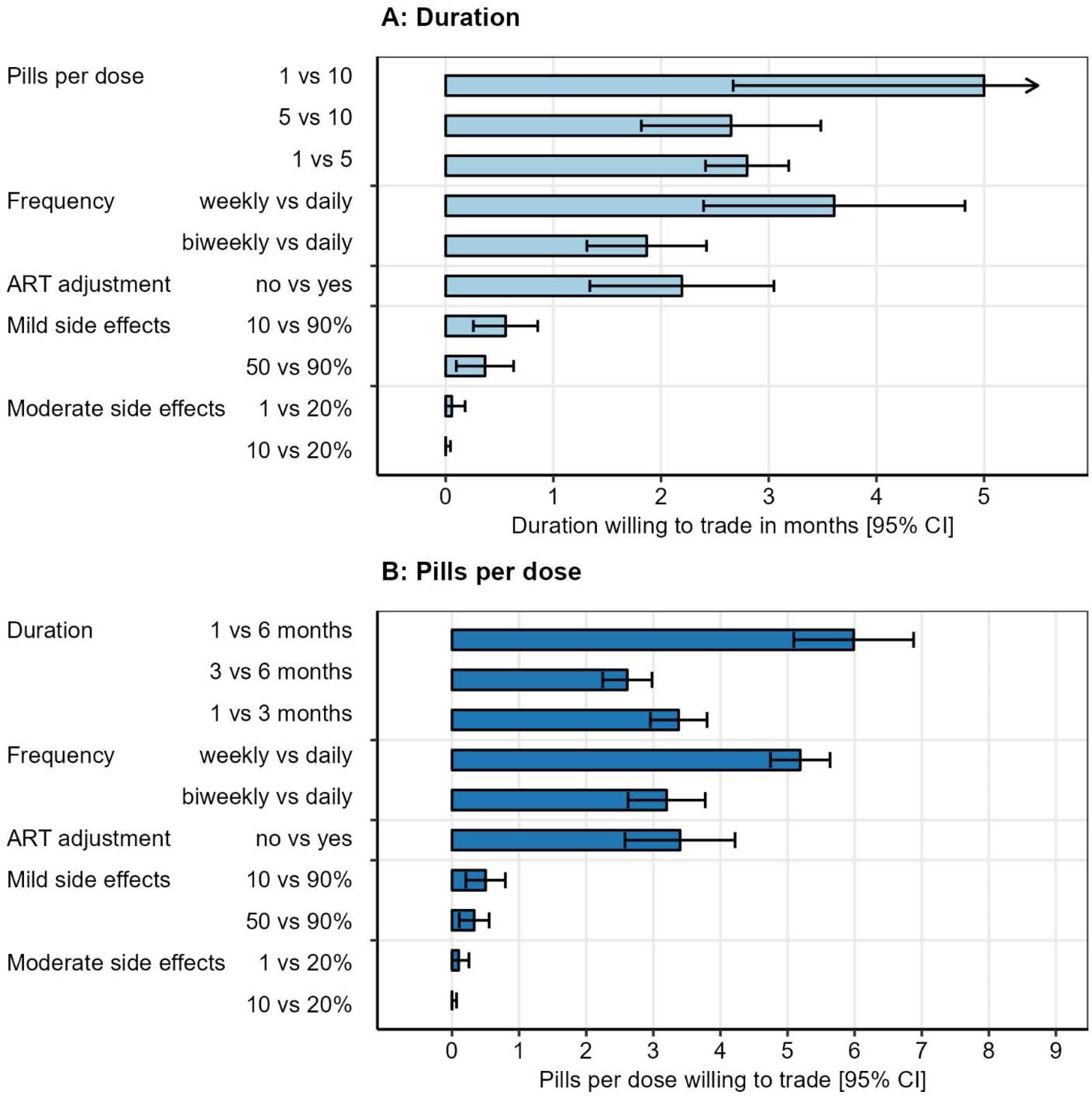
Willingness to trade (A) additional treatment duration months or (B) additional pills per dose for other improved regimen features. Results are truncated below zero months and above 5 months, and below zero pills and above 9 pills per dose (extrapolated values). The arrow in (A) indicates the upper confidence limit for 1 vs 10 pills was out of range. ART dosage adjustment was presented as requiring a second daily dose of ART. Moderate or severe side effects were described as side effects requiring medical care. ART: antiretroviral therapy

## Discussion

This DCE among adult PLHIV in Kampala, Uganda, provides important insights about what features of TPT regimens patients value the most. Although there was substantial heterogeneity of preferences as indicated by three distinct preference groups, all groups showed very strong preference for lower pill burden. While TPT regimens as short as 1 month are now available, participants were willing to accept TPT regimens approximately 3 months longer in order to take 4 fewer pills per dose, 4 months longer to have weekly rather than daily dosing and 2 months longer to avoid ART dosage adjustment. Scale-up of current regimens and future regiment development should consider pill burden, dosing frequency and compatibility with ART rather than focus exclusively on treatment duration.

Previous studies have also suggested that pill burden, dosing frequency and compatibility with ART are important considerations for TPT regimens. A study to characterize and understand gaps in the TPT care cascade among PLHIV in Uganda found that pill burden was an important barrier for patients [18]. We previously reported that 81% of PLHIV expressed a preference for 3HP over 1HP [19], supporting our finding here that less frequent dosing is preferred. Similarly, two previous studies focusing on pediatric TPT preferences in Eswatini and Peru found that less frequent dosing was valued [20,21] even though daily dosing may be easier to remember [22]. Our DCE confirms weekly dosing is preferred among adults, too, and adds nuance by demonstrating how PLHIV make trade-offs between these features and how trade-offs differ between preference subgroups.

Notably, more than one-in-four participants (“keep ART as is” group) expressed strong for maintaining their current ART regimen without adjustments. Participants with less ART experience were particularly averse to dosage changes. Additionally, those taking other medications unrelated to HIV were also more resistant changes in their ART dosage, possibly due to concerns about potential drug-drug interactions. Since 2019, the WHO has recommended dolutegravir-based ART regimens as first-line therapy for all PLHIV, and currently over 20 million PLHIV globally are receiving these regimens [23]. While no adjustments to standard daily dolutegravir dosing are recommended for 3HP [24], preliminary data suggest that an adjustment to twice daily dolutegravir dosing is likely necessary for 1HP [25]. Our findings suggest that a significant subset of PLHIV may find the tradeoff of adjusting their ART to safely accommodate 1HP (as well as 3HR and 4H) unpalatable, potentially leading to decreased acceptance of TPT if only these regimens were offered.

Participants in our study generally placed a low value on avoiding mild and moderate or severe side effects compared to other TPT features, a finding that aligns with a best-worst scaling (BWS) choice exercise among PLHIV in South Africa [26]. We also found an association between higher education levels and greater willingness to accept a high risk of mild side effects (90%), corroborating a qualitative study from South Africa that highlighted the role of education in shaping perceptions of TPT risks and benefits [27]. This underscores the importance of using culturally tailored, patient-friendly educational materials in counseling, as we did prior to administering our DCE [28], to help especially those with lower health literacy grasp the trade-offs involved in TPT acceptance. Our findings contrast with a DCE conducted among individuals with latent TB infection in Canada, where liver damage concerns related to TPT were prominent [29]. Differences in study populations, prior TPT experience, and TB risk may explain these opposing findings. For example, most of our participants (91%) had prior TPT experience, and only 24% reported experiencing any side effects.

Our study had several strengths, including an iterative DCE design process with pilot testing [30], a large and representative sample of PLHIV in care in Kampala, Uganda, and the application of latent class analysis to uncover preference heterogeneity [31]. However, our study does have some limitations. Only 9% of participants in our final sample had never taken TPT, although we found no association between prior TPT experience or regimen type (3HP or 6H) and preferences. Second, participants with low educational levels and limited health literacy might have had difficulty understanding the risks and hypothetical choices involved in the DCE. However, most participants reported the DCE was easy to understand (88%) and pilot testing had confirmed the DCE’s relevance and comprehensibility. Finally, DCEs present hypothetical choices (‘stated preferences’) that may differ from real-world decisions (‘revealed preferences’). However, their predictive value for actual health choices has been validated [8]. Moreover, they offer advantages over ‘revealed preferences,’ which are limited to existing options and cannot predict the acceptability of future TPT regimens [29].

In conclusion, our study shows that while there are heterogeneous preferences for TPT-related features among PLHIV in Uganda, there is a strong preference for regimens with lower pill burdens, less-frequent dosing, and no need for ART regimen adjustments. Most participants exhibited a willingness to undergo longer TPT regimens if they could access a TPT regimen with these preferred features. Collectively, our findings suggest that, in order to align with the preferences of PLHIV, policymakers should prioritize the implementation of fixed-dose combinations (FDC) of existing TPT regimens and that future TPT regimens should prioritize reducing pill burden over further reducing treatment duration.

## Data Availability

All data produced in the present study are available upon reasonable request to the authors.

---

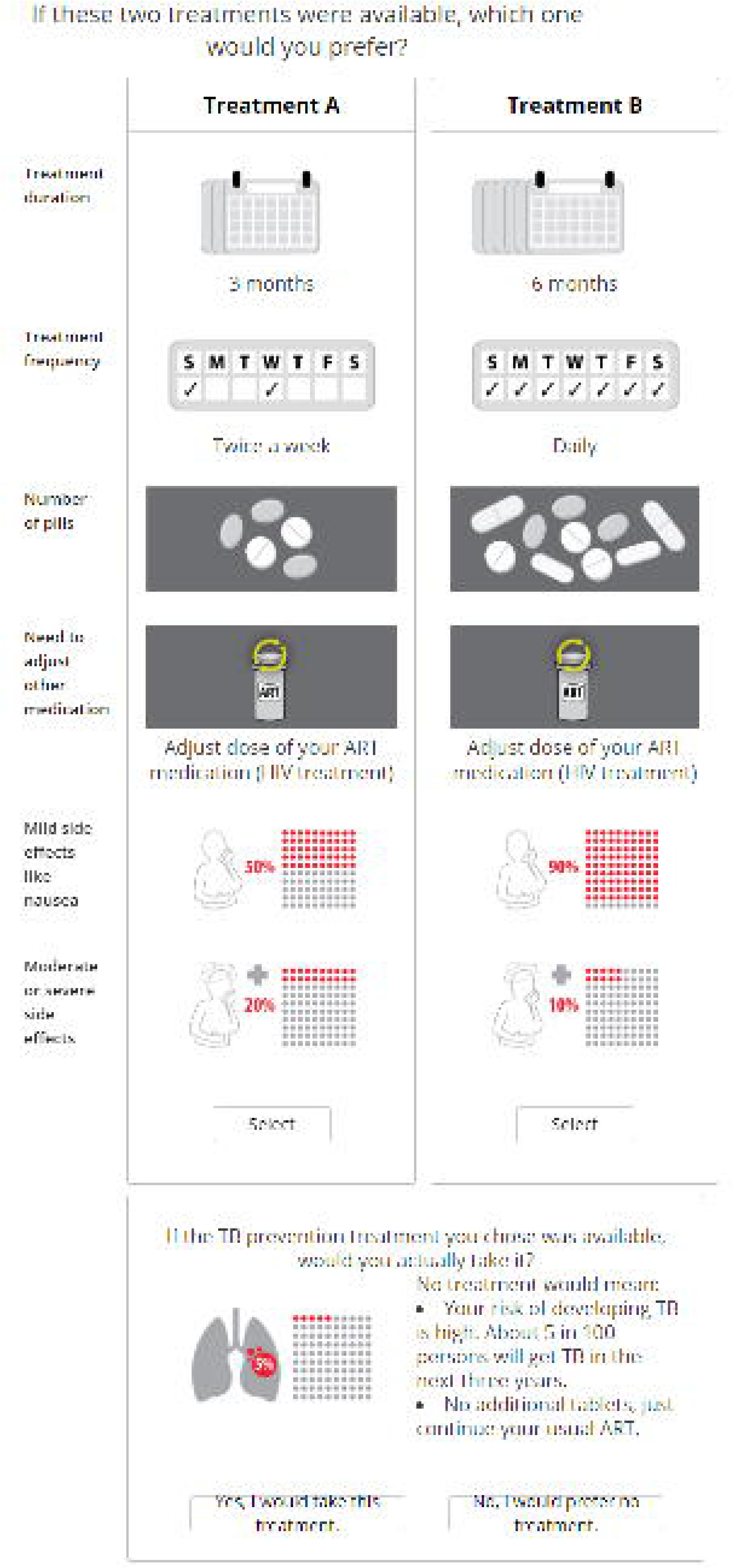

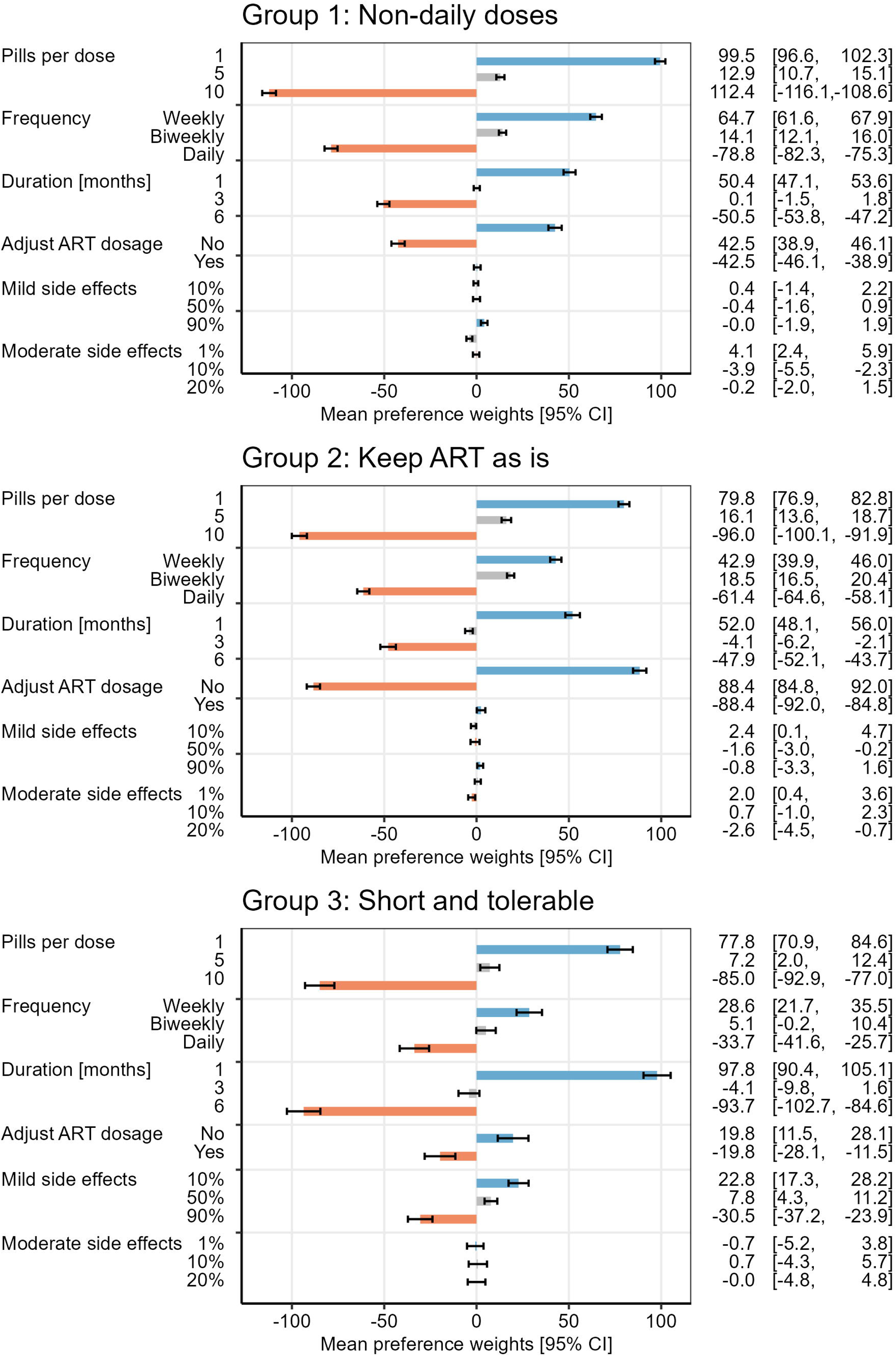

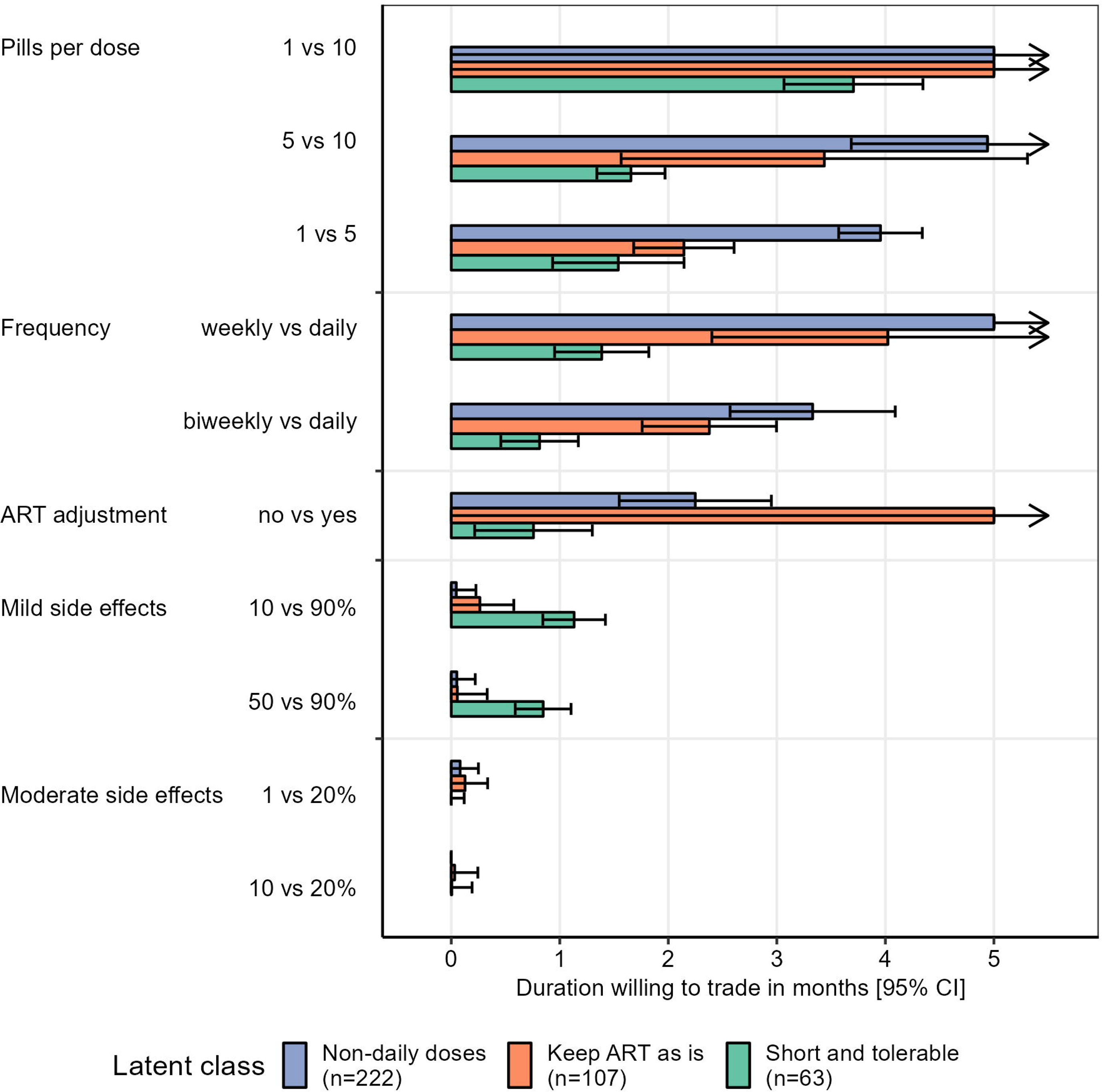

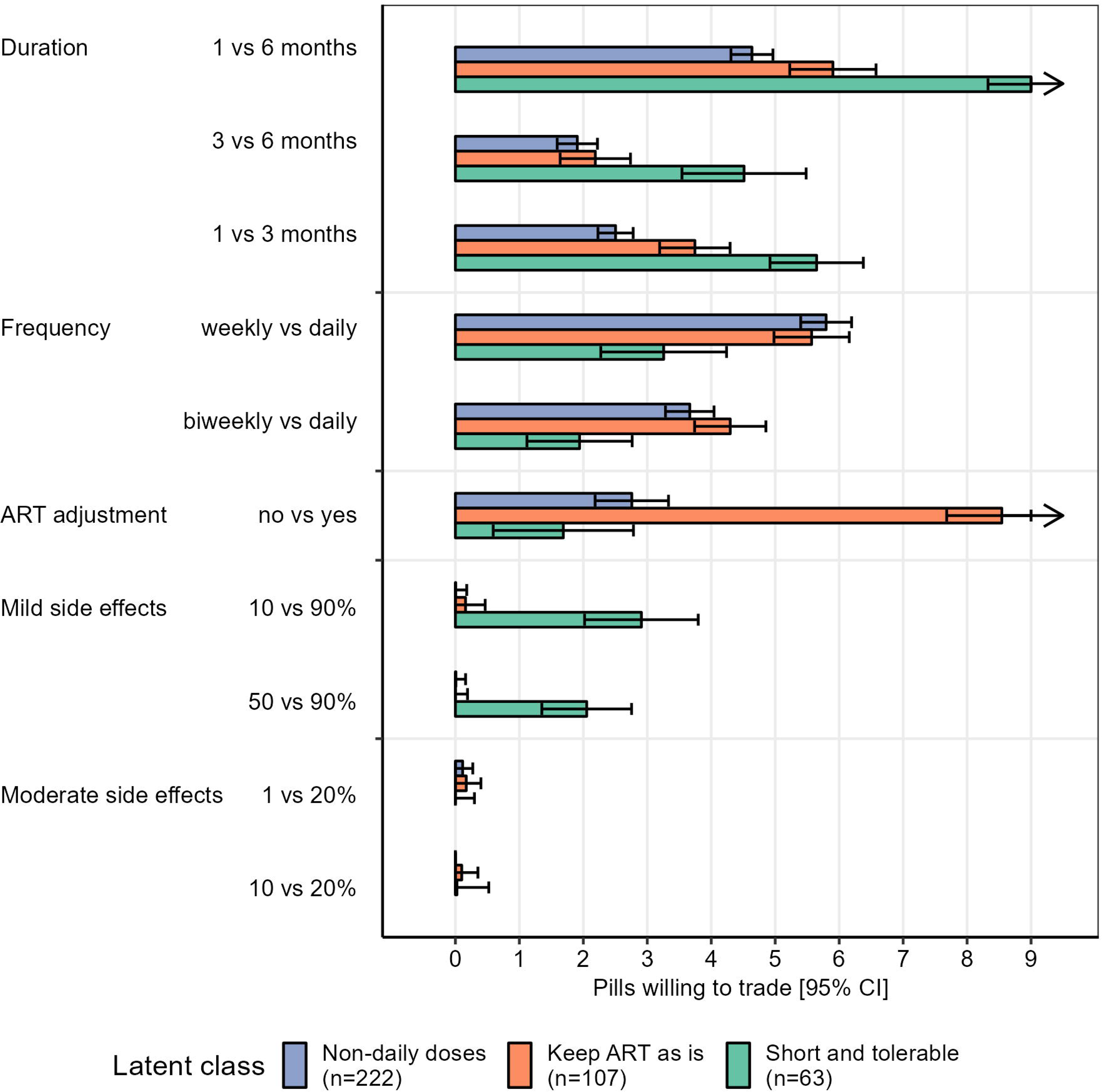

