## Appendix Table for "Preferences of people living with HIV for features of tuberculosis preventive treatment regimens – a discrete choice experiment"

**Appendix Table 1**: Levels for dominant choice task

| | **Treatment A** | **Treatment B** |
|---|---|---|
| **Duration** | 6 months | 1 month |
| **Frequency** | Daily | Daily |
| **Number of pills** | 10 | 1 |
| **Adjust ART dosage** | No | No |
| **Mild side effects** | 90% | 10% |
| **Moderate side effects** | 20% | 1% |

**Appendix Table 2**: Hierarchical bayes mean utility (392 participants). This table shows the underlying values displayed in figure 3 in the main manuscript.

| **Attribute** | **Relative importance [95% CI]** | **Level** | **Mean preference weight [95% CI]** | | |
|---|---|---|---|---|---|
| Number of pills | 32.4 [31.6, 33.2] | 1 | 90.6 | [88.3, | 92.9] |
| | | 5 | 12.9 | [11.2, | 14.5] |
| | | 10 | - | [-106.4, | -100.6] |
| Frequency | 20.5 [19.7, 21.3] | Weekly | 53.0 | [50.3, | 55.6] |
| | | Twice per week | 13.8 | [12.3, | 15.4] |
| | | Daily | -66.8 | [-69.8, | -63.8] |
| Duration (months) | 19.5 [18.6, 20.5] | 1 | 58.4 | [55.5, | 61.4] |
| | | 3 | -1.7 | [-3.2, | -0.3] |
| | | 6 | -56.7 | [-59.8, | -53.6] |
| ART adjustment | 18.2 [17.2, 19.2] | no | 51.4 | [47.8, | 54.9] |
| | | yes | -51.4 | [-54.9, | -47.8] |
| Mild side effects | 5.0 [4.6, 5.4] | 10% | 4.5 | [2.8, | 6.2] |
| | | 50% | 0.6 | [-0.4, | 1.6] |
| | | 90% | -5.1 | [-7.1, | -3.2] |
| Moderate side | 4.4 [4.1, 4.7] | 1% | 2.8 | [1.5, | 4.1] |
| | | 10% | -1.9 | [-3.2, | -0.6] |
| | | 20% | -0.9 | [-2.2, | 0.5] |
| No treatment | - | - | - | [-147.2, | -123.2] |

**Appendix Table 3**: Linear regression of preference weights for undesirable levels by participant characteristics based on hierarchical Bayesian estimation of individual preference weight. Negative values are less desirable, whereas positive are more desirable. For each undesirable level, the relative risk is given with the 95% confidence interval in brackets. Statistically significant effects at a level of 0.05 are highlighted in bold (not adjusted for multiplicity). Participants with education (compared to none) were less averse to mild side effects. Participants with longer duration since ART initiation were less averse to adjusting the ART dose, whereas participants who were taking other medications (for other conditions) were more averse to adjusting the ART dose.

| | **Duration: 6 months** | | **Number: 10 pills** | | **Frequency: daily** | | **ART: adjust dosage** | | **Mild side effects: 90%** | | **Moderate side effects: 20%** | |
|---|---|---|---|---|---|---|---|---|---|---|---|---|
| **Intercept** | -48.4 | (-63.9, -32.9) | -102.5 | (-117.1, -87.9) | -79.3 | (-94.4, -64.3) | -47.3 | (-66.5, -28.1) | -10.7 | (-20.7, -0.72) | 5.06 | (-1.75, 11.9) |
| **Female vs male** | 0.80 | (-6.40, 8.01) | 3.41 | (-3.40, 10.2) | 1.31 | (-5.70, 8.32) | -7.66 | (-16.2, 0.89) | 3.06 | (-1.61, 7.73) | -2.17 | (-5.36, 1.02) |
| **Age per 10 years (ref: 18)** | -0.76 | (-4.52, 2.99) | -0.06 | (-3.61, 3.49) | 3.38 | (-0.27, 7.03) | -1.52 | (-6.00, 2.97) | -0.39 | (-2.82, 2.05) | -0.19 | (-1.85, 1.48) |
| **Education: any vs none** | -0.74 | (-8.19, 6.72) | -4.14 | (-11.2, 2.91) | 1.64 | (-5.61, 8.89) | -8.19 | (-16.9, 0.49) | **5.61** | **(0.77, 10.4)** | -2.29 | (-5.59, 1.01) |
| **Poor vs not poor[1]** | 5.16 | (-2.98, 13.3) | 1.65 | (-6.04, 9.34) | -2.64 | (-10.6, 5.28) | -3.83 | (-13.1, 5.44) | 1.66 | (-3.59, 6.92) | -0.25 | (-3.84, 3.34) |
| **Working vs unemployed/other[2]** | -1.82 | (-9.72, 6.08) | 4.28 | (-3.19, 11.7) | 2.71 | (-4.97, 10.4) | 0.05 | (-8.92, 9.03) | -0.62 | (-5.74, 4.49) | -1.56 | (-5.05, 1.93) |
| **Prior history of TB vs none** | 5.47 | (-2.75, 13.7) | 4.66 | (-3.11, 12.4) | -7.87 | (-15.9, 0.13) | -8.55 | (-17.9, 0.80) | 3.23 | (-2.15, 8.62) | -2.42 | (-6.09, 1.26) |
| **Years on ART (ref: 0)** | -0.65 | (-1.46, 0.15) | -0.45 | (-1.21, 0.31) | 0.11 | (-0.67, 0.89) | **1.09** | **(0.18, 2.01)** | 0.07 | (-0.45, 0.59) | 0.03 | (-0.33, 0.38) |
| **Prior TPT (ref: without side effects)** | | | | | | | | | | | | |
| **Never took TPT** | | | | | | | | | -3.45 | (-10.9, 4.01) | -2.46 | (-7.56, 2.63) |
| **Prior TPT with side effects** | | | | | | | | | -3.67 | (-8.65, 1.31) | -1.87 | (-5.27, 1.54) |
| **Other medications vs none[3]** | | | | | | | **-7.70** | **(-15.3, -0.06)** | | | | |
| **Contraceptive use vs none[4]** | | | | | | | 3.12 | (-8.07, 14.3) | | | | |
| **Herbal medicine use vs never** | | | | | | | 6.45 | (-2.18, 15.1) | | | | |

[1] Multidimensionally poor or severely poor vs not vulnerable or vulnerable. [2] Working includes hired or self-employed, unemployed/other includes unemployed, not working, and other. [3] Other medications than ART or contraceptives. [4] For contraceptive use, men were coded as non-users. ART: Antiretroviral therapy, TB: active tuberculosis, TPT: tuberculosis preventive treatment.

**Appendix Table 4**: Regression of participant characteristics vs latent class membership (outcome) (relevant predictors)

| | Group 2 vs 1 | | | | Group 3 vs 1 | | | |
|---|---|---|---|---|---|---|---|---|
| | RR | (95% CI) | | p | RR | (95% CI) | | p |
| **Female vs Male** | 0.88 | (0.50, | 1.54) | 0.64 | 0.80 | (0.41, | 1.58) | 0.52 |
| **Age per 10 years (ref: 18)** | 1.09 | (0.81, | 1.47) | 0.57 | 1.25 | (0.86, | 1.81) | 0.24 |
| **Education: any vs none** | 1.41 | (0.75, | 2.66) | 0.29 | 0.61 | (0.32, | 1.16) | 0.13 |
| **Poor vs not*:** | 1.45 | (0.80, | 2.62) | 0.22 | 0.96 | (0.44, | 2.09) | 0.91 |
| **Working vs unemployed/other** | 0.91 | (0.50, | 1.65) | 0.76 | 1.31 | (0.61, | 2.84) | 0.49 |
| **Prior TPT vs never** | 1.02 | (0.42, | 2.46) | 0.97 | 0.71 | (0.25, | 2.01) | 0.52 |
| **Prior active TB vs none** | 0.93 | (0.49, | 1.74) | 0.81 | 0.51 | (0.22, | 1.20) | 0.12 |
| **Years on ART (per 5 years, ref: 10.4)** | **0.72** | **(0.53,** | **0.98)** | **0.04** | 1.02 | (0.70, | 1.48) | 0.92 |
| **Contraceptive use vs none** | 1.12 | (0.53, | 2.38) | 0.77 | 1.62 | (0.67, | 3.88) | 0.28 |
| **Other medications vs none** | 1.48 | (0.89, | 2.45) | 0.13 | 0.68 | (0.35, | 1.32) | 0.26 |

*Multidimensionally poor or severely poor, vs not vulnerable or vulnerable.

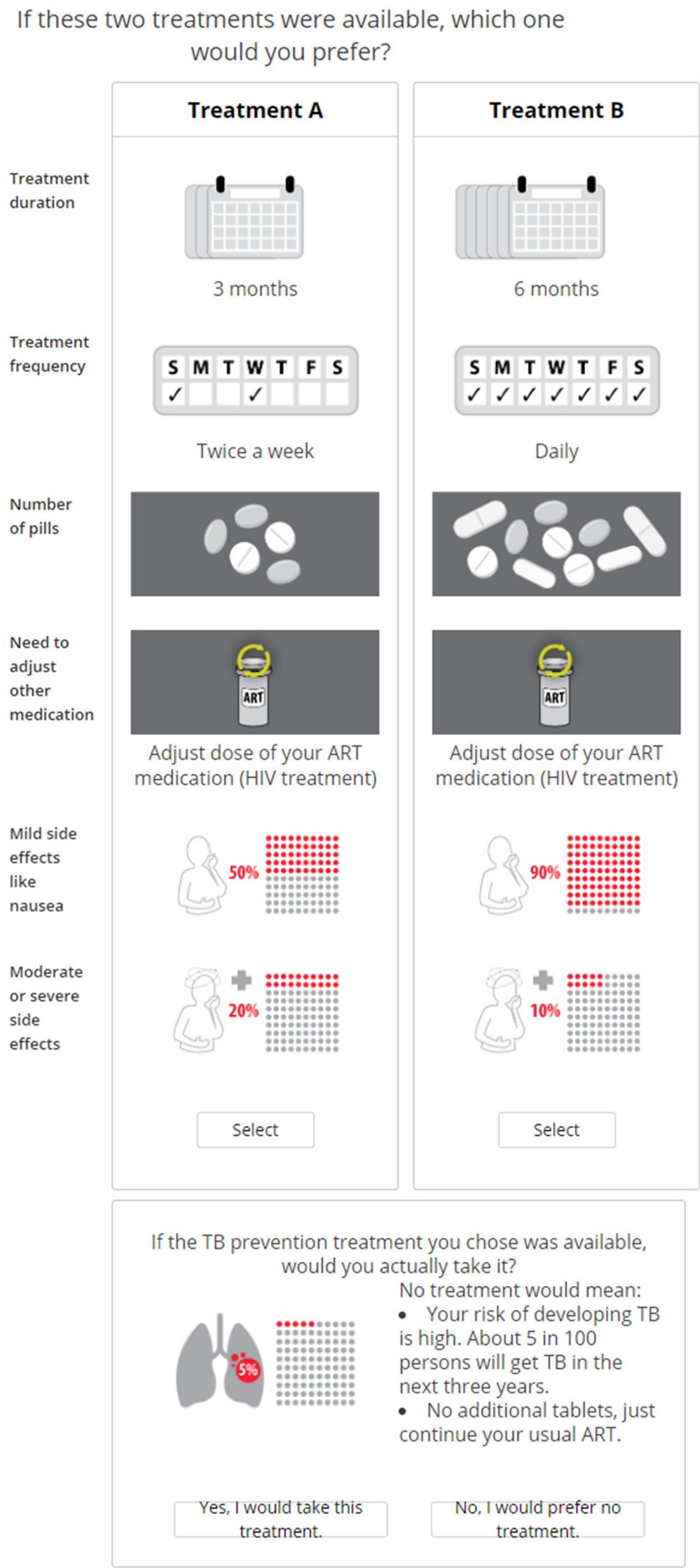


**Appendix Figure 1**: Example DCE random choice task (screenshot). The DCE was administrated on an electronic tablet. Participants were able to view all attributes and levels at once.

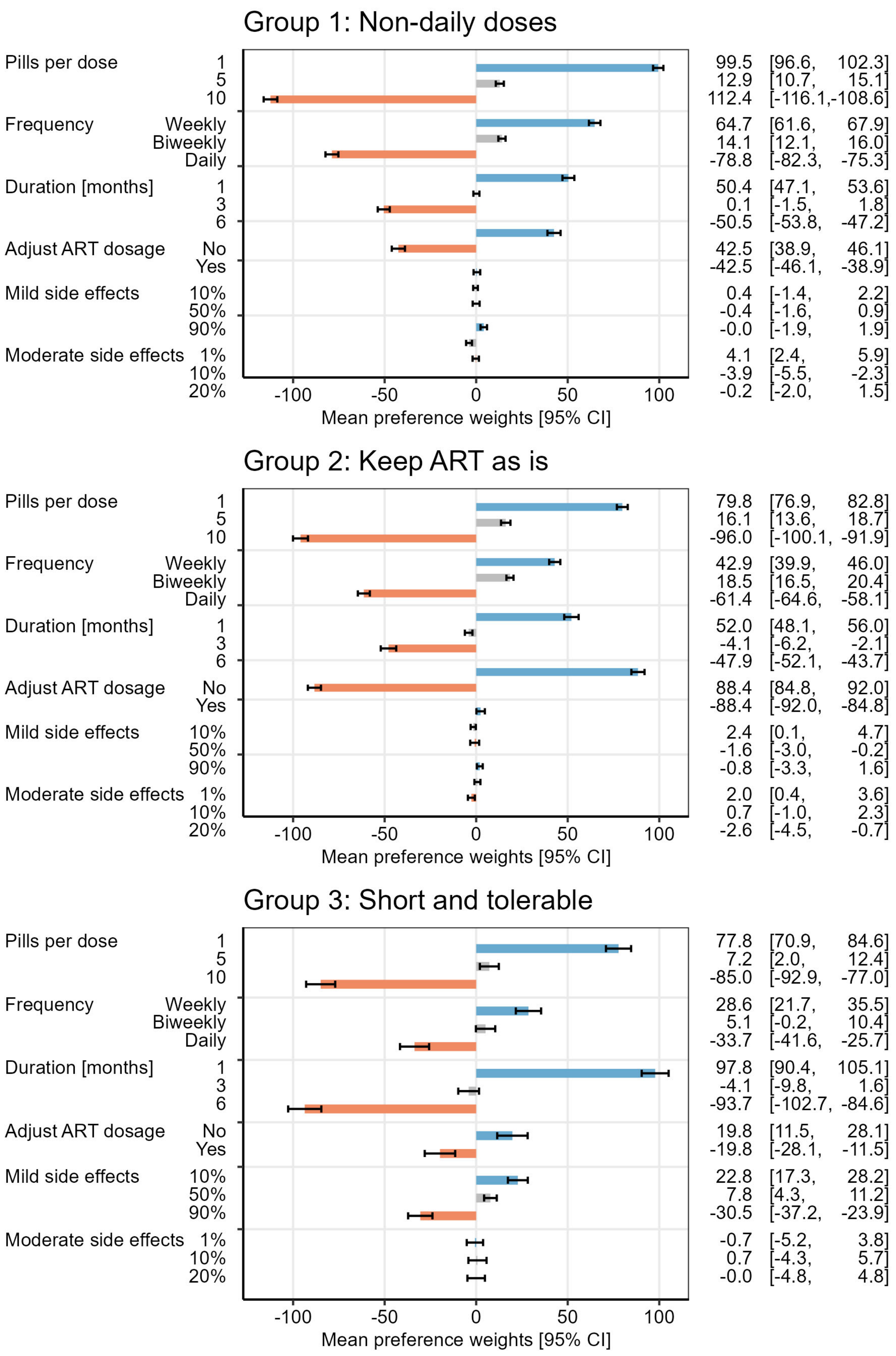


**Appendix Figure 2:** Mean preference weights by latent class [95% confidence interval]. Preference weights were estimated using hierarchical Bayes.

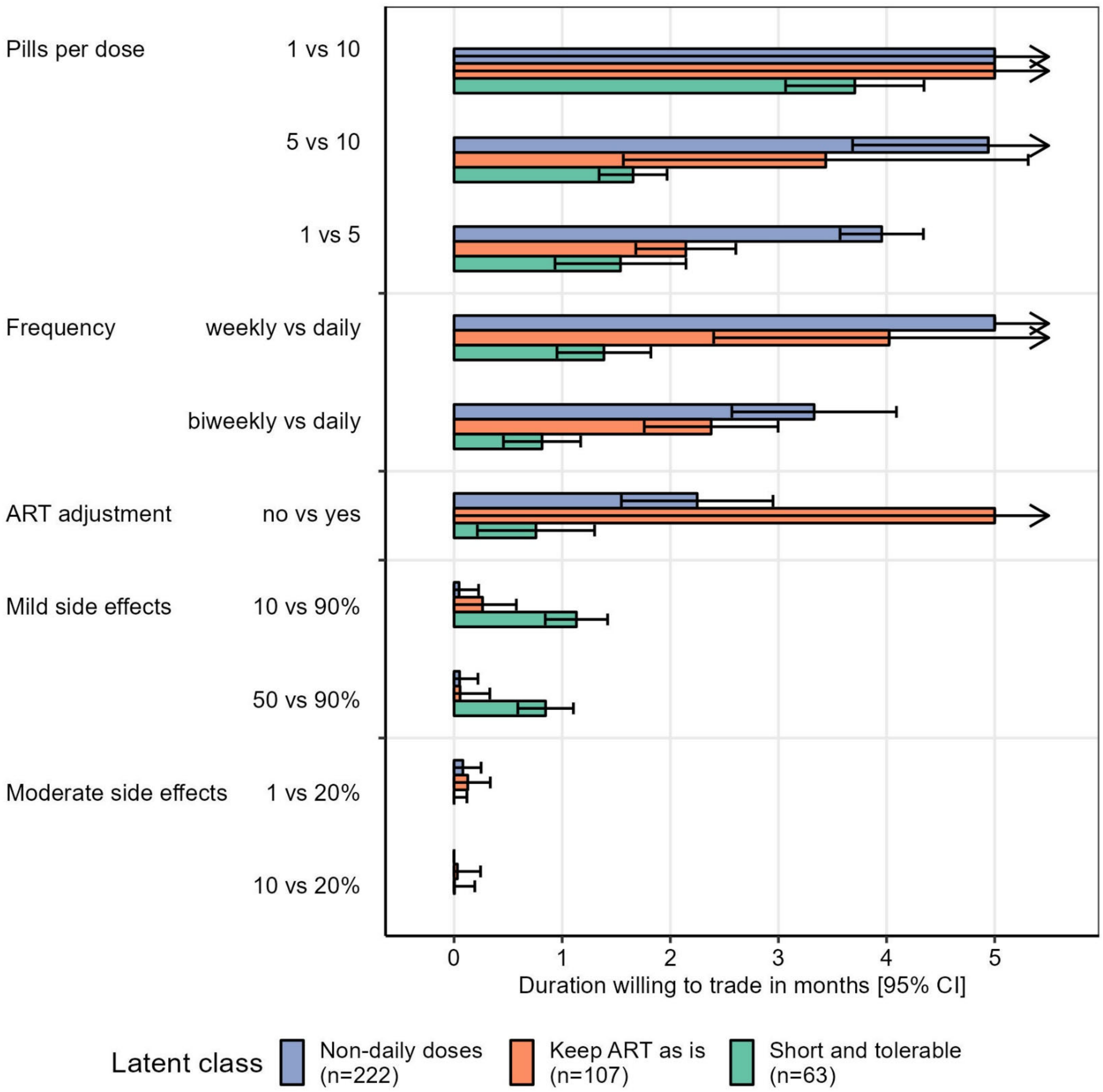


**Appendix Figure 3**: Willingness to trade longer duration with other improved treatment features by preference groups identified by latent class analysis. The simulation used two competitor products, 6 months of daily isoniazid (2 pills) and 3 months of weekly isoniazid and rifampin (5 pills), based on currently available TPT regimens in Uganda. For legibility, negative values were truncated at 0 months. Error bars show the 95% confidence interval. Arrows indicate values beyond the level range (above 5 months). Arrows without an error bar indicate that all values, including the lower bound of the confidence interval, were above 5 months (weekly versus daily frequency). Arrows with an error bar starting at zero denote estimates with wide confidence intervals from below zero to above 5 months (1 vs 10 pills and ART adjustment no vs yes).

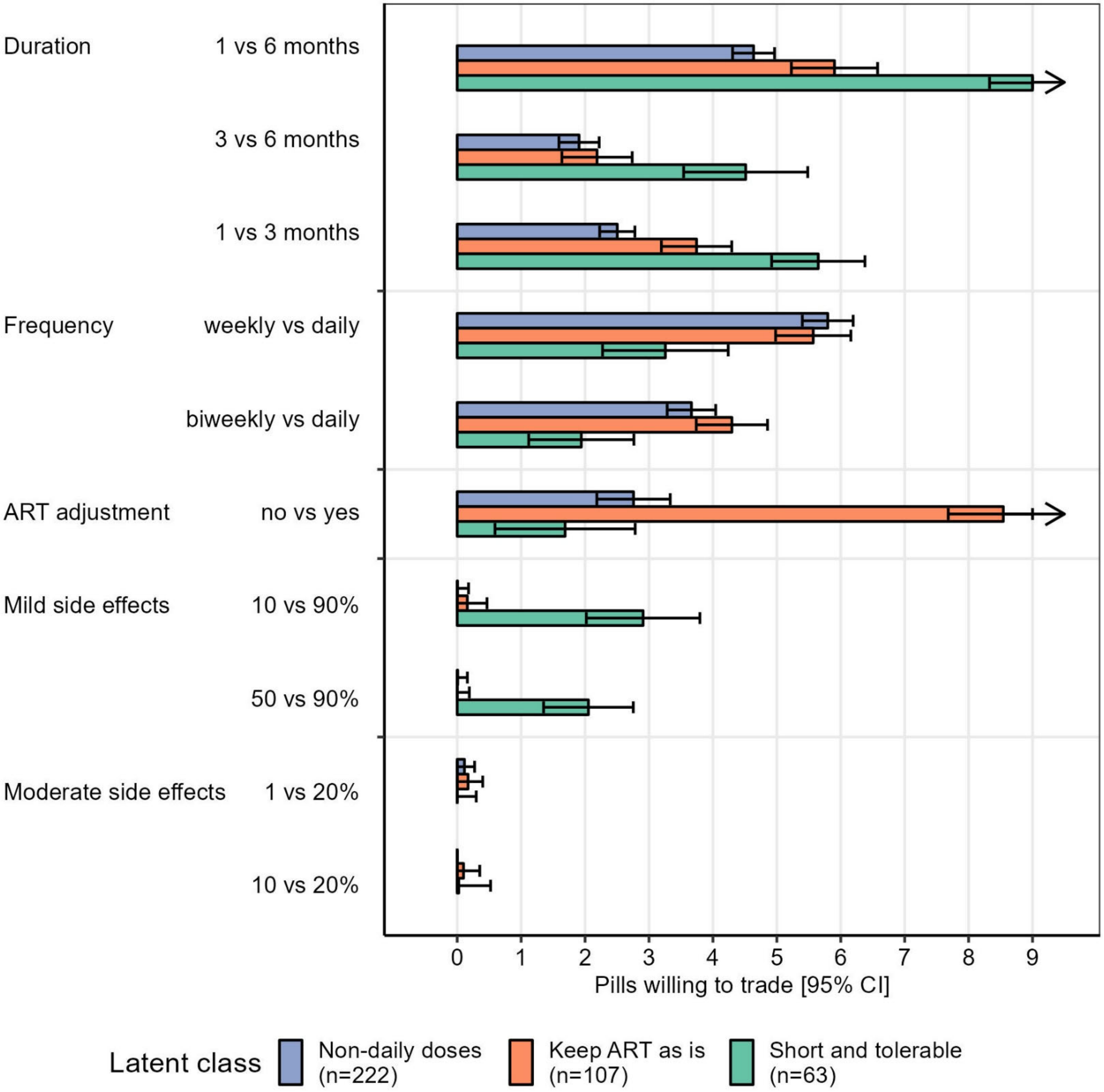


**Appendix Figure 4**: Willingness to trade additional pills with other improved treatment features by preference groups identified by latent class analysis. The simulation used two competitor products, 6 months of daily isoniazid (2 pills) and 3 months of weekly isoniazid and rifampin (5 pills), based on currently available TPT regimens in Uganda. For legibility, negative values were truncated at 0 months. Error bars show the 95% confidence interval. Arrows indicate values beyond the level range (above 9 pills).
