## Appendix A for "Preferences of people living with HIV for features of tuberculosis preventive treatment regimens – a discrete choice experiment"

| **Questions for participant in the pilot test**: |
| --- |
| 1. Was there anything in the survey that you did not understand or that was hard to understand? |
| 2. Overall, how difficult was it to understand the questions you were asked?<br>☐ Very difficult<br>☐ Difficult<br>☐ Neutral<br>☐ Easy<br>☐ Very easy |
| 3. How difficult was it for you to choose between the two different preventive treatment options in each question?<br>☐ Very difficult<br>☐ Difficult<br>☐ Neutral<br>☐ Easy<br>☐ Very easy |
| 4. How difficult was it for you to choose whether to take the selected preventive treatment or no treatment in each question?<br>☐ Very difficult<br>☐ Difficult<br>☐ Neutral<br>☐ Easy<br>☐ Very easy |
| 5. Did you have a strategy for choosing between the two preventive treatment options in each question? (select all that apply)<br>☐ I did not have a strategy<br>☐ I focused on a just a few aspects of the preventive treatment<br>☐ I considered most of the aspects of the preventive treatment<br>☐ I considered all of the aspects of the preventive treatment<br>☐ I avoided some aspects that were not acceptable to me or undesirable (e.g. high risk of side effects or high number of pills), please specify which:<br><br>☐ Other, please specify: |

6. If you considered a few or most of the aspects, which ones did you consider?

| I considered these: | I did not consider these: |
|---|---|
| ☐ Number of pills | ☐ Number of pills |
| ☐ Treatment duration | ☐ Treatment duration |
| ☐ Treatment frequency | ☐ Treatment frequency |
| ☐ Effects on other medications | ☐ Effects on other medications |
| ☐ Mild side effects | ☐ Mild side effects |
| ☐ Moderate or severe side effects | ☐ Moderate or severe side effects |
| ☐ Tuberculosis risk | ☐ Tuberculosis risk |

7. Thinking back to when you had two choose between two preventive treatments. Is there anything more you can tell me about how you made your decisions?
   Use the following prompts if needed: Did you develop some kind of system? Which features were most important to you? Were there features that you did not care about and ignored when comparing treatments? Were there any particularly undesirable features that you avoided?

8. Can you explain in your own words how the tuberculosis risk varied between treatments? And how did the TB risk vary between taking a treatment and no treatment?

9. Does this difference matter to you?

10. Can you explain in your own words how treatments differed in their possible effect on other treatments?

11. Does this matter to you? Why?

12. How did you feel about the survey in general?

13. How did you feel about the length of the survey?

14. Were the graphics clear?

15. Was the screen size of the tablet and the font size ok?

16. Do you have any concerns about having a facilitator from the study team help you with the survey?

17. Is there anything else you think could be improved?

| **Questions for the study team:** |
| --- |
| Any comments on the procedures? What could be improved? |
| Any comments based on your observation:<br>Did the participant appear to understand the tasks?<br>Did the participant focus on any specific features in the DCE, or ignore other features? E.g. did the participant ask you to repeat any difference between features, or mention that they would not care about a feature?<br>How long did the participant stay motivated and focused? E.g. did the participant appear to slow down or get fatigued at any point during the survey? Please explain. |
