## Appendix B for "Preferences of people living with HIV for features of tuberculosis preventive treatment regimens – a discrete choice experiment"

Appendix B – DCE education flipbook: Preferences of people living with HIV for features of tuberculosis preventive treatment regimens – a discrete choice experiment

Hélène E. Aschmann, Allan Musinguzi, Jillian L. Kadota, Catherine Namale, Juliet Kakeeto, Jane Nakimuli, Lydia Akello, Fred Welishe, Anne Nakitende, Christopher Berger, David W. Dowdy, Adithya Cattamanchi, Fred C. Semitala, Andrew D. Kerkhoff

### Discrete Choice Experiment (DCE): Patient Education

### Features of different treatments

1

- We will ask you questions about your preferences for potential future options of taking tuberculosis preventive therapy.
- When we ask you these questions about your preferences, we will present different scenarios to you, and we will ask you to choose which one you would prefer.
- Here we explain the features of different treatments.

2

#### Treatment duration

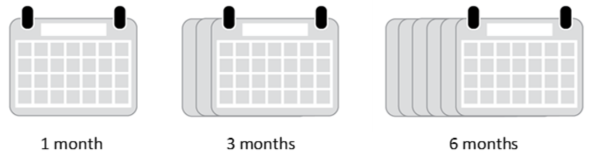


3

**Treatment duration**

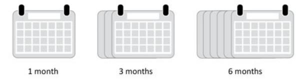


- Different treatments for preventing active TB exist, and they differ in duration.
- The treatments we will show you range from 1 month to 6 months.

4

### Treatment frequency

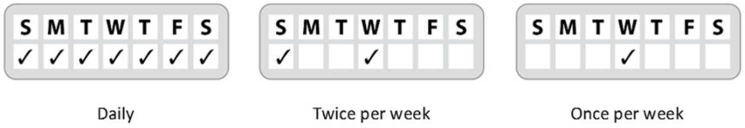


5

#### Treatment frequency

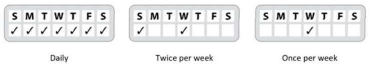


- Some treatments require daily pills, others require pills two times per week, and others are only once weekly

6

### Number of pills

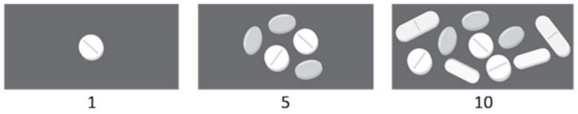


7

#### Number of pills

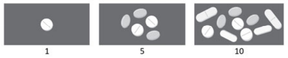


- Some treatments consist of only one tablet per dose, but some may require you to take up to 10 tablets each dose.
- These tablets would be additional tablets on top of your antiretroviral treatment.

8

#### Interaction with other drugs

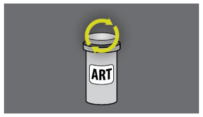


Adjust the dose of
your ARV medication.

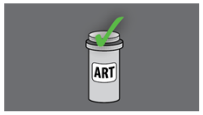


No dose adjustment needed.
You can keep taking your ARV.

9

On these treatments, you will need to adjust the dose (e.g. one more pill a day) of your **ARV medication** (medication for HIV).

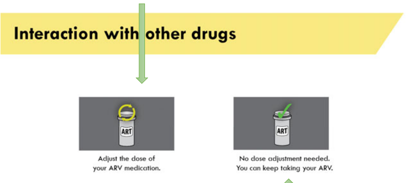


These treatments do not affect your ARV. You can keep taking the ARV medication you are taking now.

- Some treatments affect how other drugs work. For example, some TB preventive therapies require people to increase the dose of their ARV treatment. This would mean taking your ARVs twice daily during your TB preventive therapy. You would take an extra pill of the same ARV drugs.
- For these hypothetical treatment choices, the icons indicate which treatments may need adjustments to ARV treatment. Please think about whether this would matter to you.

10

#### Short-lasting, mild side effects

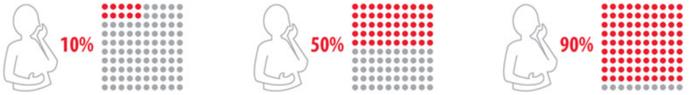


10 in 100 persons experience mild side effects

50 in 100 persons experience mild side effects

90 in 100 persons experience mild side effects

11

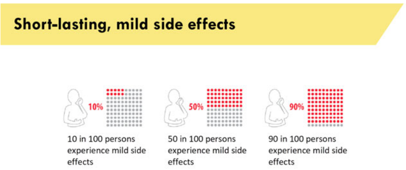


- Some TB preventive therapies lead to mild side effects such as nausea, dizziness, fatigue, or skin rashes for a week or so. These symptoms may bother you and may impact your activities, but after you wait a few days, they disappear. With mild side effects like these, you typically do not stop the medication, and you do not need to see your doctor.
- Different treatments differ in how frequently mild side effects occur.

12

#### Moderate or severe side effects

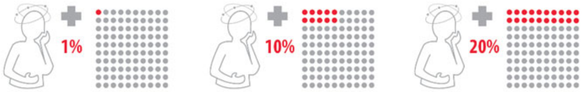


1 in 100 persons experience more severe side effects

10 in 100 persons experience more severe side effects

20 in 100 persons experience more severe side effects

13

##### Moderate or severe side effects

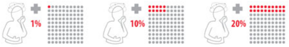

1 in 100 persons experience more severe side effects

10 in 100 persons experience more severe side effects

20 in 100 persons experience more severe side effects

- Some people may develop more severe symptoms that do not quickly go away on their own and may be very bothersome.
- These side effects include tingling feet, allergy, or liver damage.
- These typically require additional visits to the healthcare facility and may require you to stop your medicine or be admitted to the hospital.
- Different treatments differ in how frequently moderate or severe side effects occur.

14

### Choice tasks

15

16

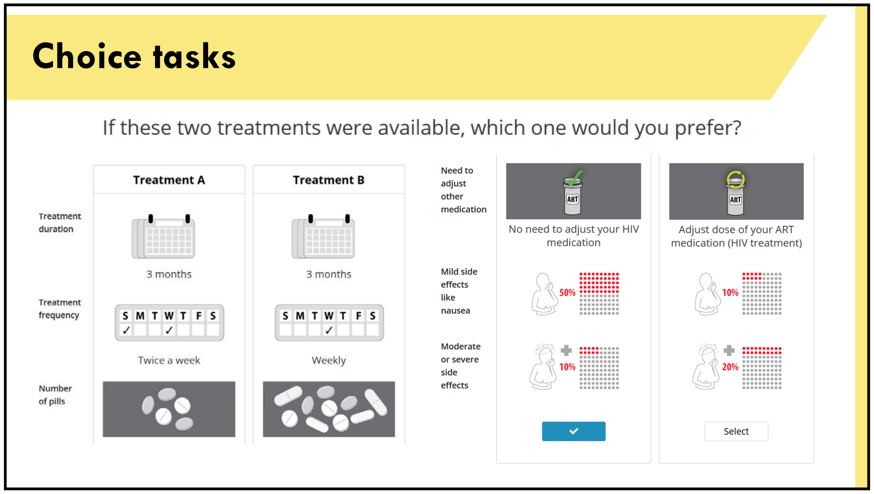


17

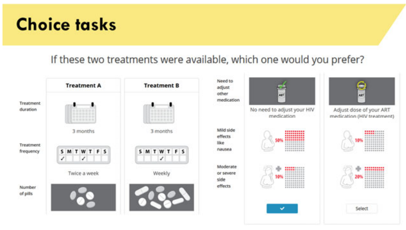


- We will now ask you to repeatedly make a choice between two treatments that may exist in the future. This is how the task could look like:
- Each treatment will show different combinations for the six features.
- In this example, the respondent would have chosen treatment A.

18

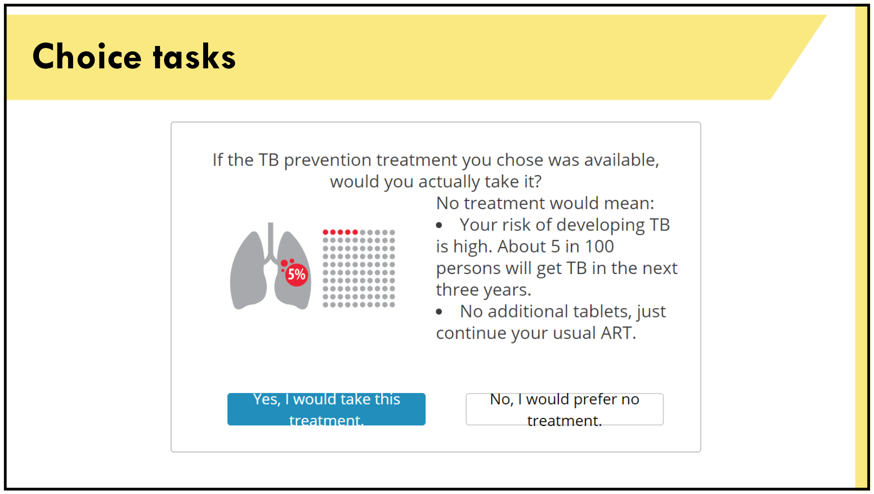


19

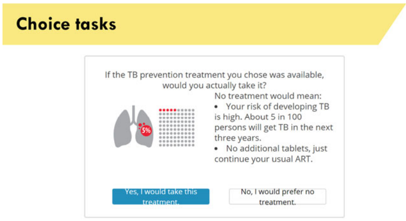


- Now we would like to know if the respondent would actually take treatment A or prefer no treatment.
- In our hypothetical example, if the respondent does not take treatment, the risk to develop TB in the next 3 years is 5%. In other words, 5 in 100 persons will develop active TB. If the respondent takes treatment A, the risk is smaller, about half as large.
- No treatment means no additional tablets and no side effects, the patient just continues the normal ARV treatment.
- In this example, the respondent answered that they would take the treatment.

**Now it is your turn!**

**We will show you hypothetical treatments and want to know how you feel about them.**

20

#### Questions?

Do you have any questions about these choice tasks?

21

Questions?

Do you have any questions about these choice taks?

**INSTRUCTIONS FOR STUDY STAFF:**

- Make sure you answer any participant questions or concerns.
- The participant is allowed to go back to the explanations of the features and of the tasks at any time during the survey.

22
